# Anti-nucleocapsid antibodies following SARS-CoV-2 infection in the blinded phase of the mRNA-1273 Covid-19 vaccine efficacy clinical trial

**DOI:** 10.1101/2022.04.18.22271936

**Authors:** Dean Follmann, Holly E. Janes, Olive D. Buhule, Honghong Zhou, Bethany Girard, Kristen Marks, Karen Kotloff, Michaël Desjardins, Lawrence Corey, Kathleen M. Neuzil, Jacqueline M. Miller, Hana M. El Sahly, Lindsey R. Baden

## Abstract

**Importance:** The performance of immunoassays for determining past SARS-CoV-2 infection, which were developed in unvaccinated individuals, has not been assessed in vaccinated individuals.

**Objective:** To evaluate anti-nucleocapsid antibody (anti-N Ab) seropositivity in mRNA-1273 vaccine efficacy trial participants after SARS-CoV-2 infection during the trial’s blinded phase.

**Design:** Nested analysis in a Phase 3 randomized, placebo-controlled vaccine efficacy trial. Nasopharyngeal swabs for SARS-CoV-2 PCR testing were taken from all participants on Day 1 and Day 29 (vaccination days), and during symptom-prompted illness visits. Serum samples from Days 1, 29, 57, and the Participant Decision Visit (PDV, when participants were informed of treatment assignment, median day 149) were tested for anti-N Abs.

**Setting:** Multicenter, randomized, double-blind, placebo-controlled trial at 99 sites in the US.

**Participants:** Trial participants were ≥ 18 years old with no known history of SARS-CoV-2 infection and at appreciable risk of SARS-CoV-2 infection and/or high risk of severe Covid-19. Nested sub-study consists of participants with SARS-CoV-2 infection during the blinded phase of the trial.

**Intervention:** Two mRNA-1273 (Moderna) or Placebo injections, 28 days apart.

**Main Outcome and Measure:** Detection of serum anti-N Abs by the Elecsys (Roche) immunoassay in samples taken at the PDV from participants with SARS-CoV-2 infection during the blinded phase. The hypothesis tested was that mRNA-1273 recipients have different anti-N Ab seroconversion and/or seroreversion profiles after SARS-CoV-2 infection, compared to placebo recipients. The hypothesis was formed during data collection; all main analyses were pre-specified before being conducted.

**Results:** We analyzed data from 1,789 participants (1,298 placebo recipients and 491 vaccine recipients) with SARS-CoV-2 infection during the blinded phase (through March 2021). Among participants with PCR-confirmed Covid-19 illness, seroconversion to anti-N Abs at a median follow up of 53 days post diagnosis occurred in 21/52 (40%) of the mRNA-1273 vaccine recipients vs. 605/648 (93%) of the placebo recipients (p < 0.001). Higher SARS-CoV-2 viral copies at diagnosis was associated with a higher likelihood of anti-N Ab seropositivity (odds ratio 1.90 per 1-log increase; 95% confidence interval 1.59, 2.28).

**Conclusions and Relevance:** As a marker of recent infection, anti-N Abs may have lower sensitivity in mRNA-1273-vaccinated persons who become infected. Vaccination status should be considered when interpreting seroprevalence and seropositivity data based solely on anti-N Ab testing

**Trial Registration:** ClinicalTrials.gov NCT04470427

**Key Points:** *Question:* Does prior mRNA-1273 vaccination influence anti-nucleocapsid antibody seroconversion and/or seroreversion after SARS-CoV-2 infection?

*Findings:* Among participants in the mRNA-1273 vaccine efficacy trial with PCR-confirmed Covid-19, anti-nucleocapsid antibody seroconversion at the time of study unblinding (median 53 days post diagnosis and 149 days post enrollment) occurred in 40% of the mRNA-1273 vaccine recipients vs. 93% of the placebo recipients, a significant difference. Higher SARS-CoV-2 viral copy number upon diagnosis was associated with a greater chance of anti-nucleocapsid antibody seropositivity (odds ratio 1.90 per 1-log increase; 95% confidence interval 1.59, 2.28). All infections analyzed occurred prior to the circulation of delta and omicron viral variants.

*Meaning:* Conclusions about the prevalence and incidence of SARS-CoV-2 infection in vaccinated persons based on anti-nucleocapsid antibody assays need to be weighed in the context of these results.

## Introduction

A reliable diagnostic tool to identify recent and remote infection with SARS-CoV-2 is needed to enable population-based seroprevalence studies, to ascertain infection status in vaccine clinical trials, and to diagnose post-infectious complications of Covid-19 such as the multisystem inflammatory syndrome. Antibodies against the nucleocapsid protein (anti-N Abs) are not elicited by Covid-19 vaccines that target the spike protein such as all vaccines used in the US, and assays that measure anti-N Abs have demonstrated high sensitivity and specificity in many studies when used at a minimum of 14 days post infection.^1^ In addition, the anti-N Ab response in unvaccinated persons has been reported to be durable, with half-life estimates ranging from 68 to 283 days.^2,3^ The determination of previous SARS-CoV-2 infections in participants in the pivotal Phase 3 clinical trials^4,5^ and in seroprevalence studies conducted early in the pandemic^6^ relied on detection of anti-spike or anti-N Abs via chemiluminescence immunoassays in unvaccinated persons. However, it is unknown how vaccine induced immunity may impact seroconversion to non-Spike proteins, and whether such immunoassays have similar performance in determining previous SARS-CoV-2 infection in regions of the world with high vaccination coverage. To answer this question, we evaluated vaccine and placebo recipients in the mRNA-1273 Phase 3 clinical trial^4,5^ in whom SARS-CoV-2 infection was detected during the blinded placebo-controlled phase of the trial. Serum samples from these participants were assayed for anti-N Abs through ∼5 months post-enrollment.

## Methods

### Study Design and Population

Between July 27 and October 23, 2020, the COVE trial enrolled 30,420 US adults ≥ 18 years of age at appreciable risk of SARS-CoV-2 infection and/or high risk of severe Covid-19. Participants were randomized 1:1 to two 100 μg doses of mRNA-1273 vaccine or placebo, given at Day 1 (baseline) and Day 29. Participant characteristics, study procedures, and primary efficacy results are described elsewhere.^4,5^ SARS-CoV-2 infections were detected both based on symptom-prompted testing and planned diagnostic testing at specific study visits (see Figure 1). Blood was collected for anti-N Ab serology at Day 1, Day 29, Day 57, and at the Participant Decision Visit (PDV), when participants were informed about their randomization assignment and offered mRNA-1273 vaccination if previously assigned to placebo. Nasopharyngeal (NP) swabs for SARS-CoV-2 RT-PCR testing were collected at Day 1, Day 29, and the PDV in all participants. Participant PDVs occurred a median of 149 days after enrollment (IQR: 133 to 164 days). In addition, participants meeting predefined clinical criteria for suspected Covid-19 during follow-up underwent an illness visit for collection of a NP swab for SARS-CoV-2 PCR testing (nasal swabs or saliva samples could also be submitted if a clinic or home visit could not be completed). This analysis is based on data through the completion of the blinded phase of the study with a data cutoff date of March 26, 2021. eFigure 1 depicts the flow of participants with detected SARS-CoV-2 infection from enrollment through to inclusion in the analysis.

**Figure 1.**
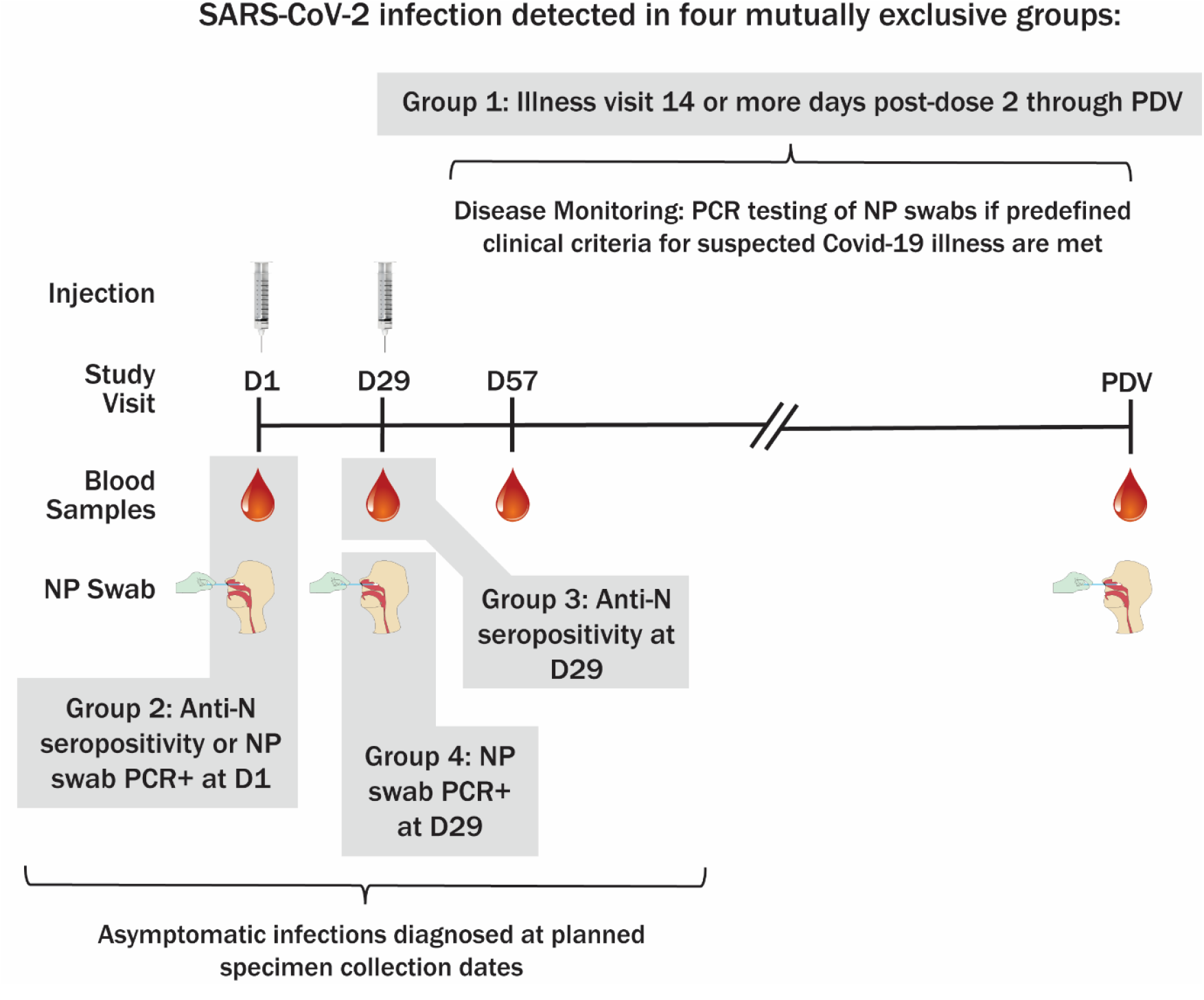
Method of SARS-CoV-2 infection determination and sampling schedule. PDV, participant decision visit.

### Laboratory Assays

Anti-N Abs were detected using the Roche Elecsys Anti-SARS-CoV-2 immunoassay (Roche Diagnostics International)^7^ on samples from Day 1, 29, 57 and the PDV. The Elecsys assay has a binary interpretation, “non-reactive” or “reactive” for the presence of anti-N Abs. Day 57 50% inhibitory dilution (ID50) neutralizing antibody titers, measured by an assay using SARS-CoV-2 spike-pseudotyped virus, were previously reported.^8^ NP swabs, nasal swabs, and saliva samples were tested for SARS-CoV-2 via RT-PCR (Viracor, Eurofins Clinical Diagnostics). In Covid-19 cases, viral copy number was assessed at the illness visits Day 1, 3, 5, 7, 9, 14, 21, and 28 by SARS-CoV-2 RT–qPCR and conversion of cycle-threshold (Ct) values to viral genome copy number.^9^ Illness visit day 1 was by swab and other illness days by saliva.

### Definitions

This study compares the outcome of anti-N seropositivity at the PDV between mRNA-1273 vaccine vs. placebo recipients with SARS-CoV-2 infection detected prior to the PDV in the Full Analysis set of randomized participants who received at least one dose of vaccine or placebo. The analysis is stratified by method and timing of SARS-CoV-2 infection detection; four mutually-exclusive groups of infections are defined as follows. The first group of infections are primary endpoint Covid-19 cases detected at an illness visit at a minimum of 14 days post dose 2 and up to the PDV. Covid-19 cases are defined as baseline SARS-CoV-2 negative participants in the per protocol population, who had at least two of the following symptoms: fever (temperature ≥38°C), chills, myalgia, headache, sore throat, or new olfactory or taste disorder, or as occurring in those who had at least one respiratory sign or symptom (including cough, shortness of breath, or clinical or radiographic evidence of pneumonia), and at least one NP swab, nasal swab, or saliva sample (or respiratory sample, if the participant was hospitalized) that was positive for SARS-CoV-2 by RT-PCR.^4,5^ The second group of infections are those anti-N seropositive by Elecsys assay or NP positive by RT-PCR at baseline (Day 1). The third group of infections are those anti-N seropositive at Day 29 without prior evidence of infection by serology or RT-PCR. The fourth group of infections consists of those NP positive by RT-PCR at Day 29, seronegative at Day 29 without prior evidence of infection by serology or RT-PCR.

### Statistical Analyses

Analyses to assess PDV anti-N seropositivity by infection group and randomization arm were pre-specified. The outcome of PDV anti-N seropositivity was compared between infection groups and randomization arms using Fisher’s exact tests. The association of PDV anti-N seropositivity with time since Covid-19 diagnosis and other factors was assessed using logistic regression. A Wilcoxon rank sum test was used to compare anti-spike antibody responses between those anti-N seropositive and seronegative at the PDV. A Wald test of interaction was used to assess whether the effect of illness visit viral copy number on PDV anti-N seropositivity differed between randomization arms. Receiver operating characteristic curves were compared using a nonparametric approach.^10^ All tests are two-sided with type I error rate equal to 0.05 and no adjustment for multiplicity.

## Results

SARS-CoV-2 infection prior to PDV was detected in four mutually exclusive groups. Eight hundred twelve baseline SARS-CoV-2 negative participants (754 placebo recipients, 58 vaccine recipients) (eTable 1) experienced illness 14 or more days after full receipt of study vaccine or placebo and were diagnosed with a PCR swab obtained at an illness visit associated with that event. For the other three groups, SARS-CoV-2 infection was first detected through planned specimen collections at baseline or D29. Infection was first detected at baseline by anti-N seropositivity or NP swab RT-PCR positivity in 684 participants (337 placebo recipients, 347 vaccine recipients) (eTable 2). Post-baseline infections were first detected by anti-N seropositivity at Day 29 in 107 participants (64 placebo recipients, 43 vaccine recipients), and by PCR positivity at Day 29 in 49 participants (36 placebo recipients, 13 vaccine recipients) (eTable 3). For each of these four groups of participants, eTables 1-3 also present their baseline characteristics by arm.

Table 1 shows anti-N seropositivity at the PDV for participants with Covid-19 detected at an illness visit. There was a substantial between-arm difference in anti-N seropositivity at the PDV: 40.4% (21/52) in vaccine recipient Covid-19 cases vs. 93.3% (605/648) in placebo recipient Covid-19 cases (p < 0.001). The median time in days from illness to PDV was 53 days with a range of 5 to 150 days. Anti-N seropositivity at the PDV was similar when stratified by median days from illness (p>0.05). Of the 52 vaccine recipients, 36 had Day 57 ID50 anti-spike neutralizing antibody titer measured as part of the case-cohort immunogenicity and immune correlates assessment^8^ and also serostatus measured at the PDV. Among these 36 individuals, the median Day 57 anti-spike ID50 titers were not significantly different between the 20 who were anti-N seronegative vs. the 16 who were anti-N seropositive at PDV (200 vs. 158 IU50/ml; p>0.05).

**Table 1.**
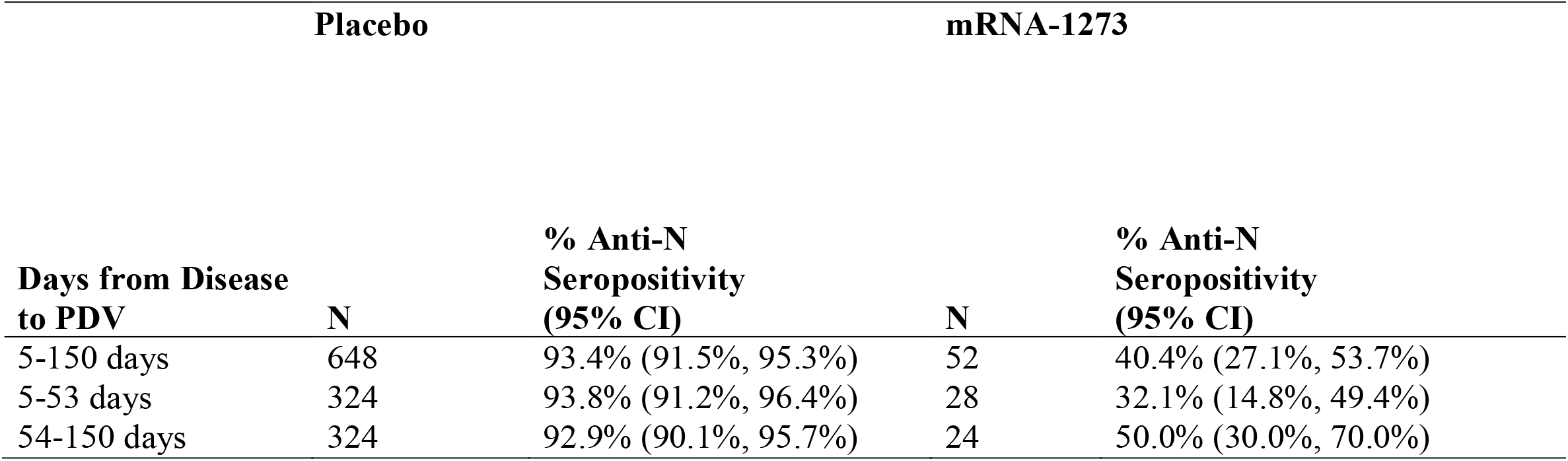
Anti-N seropositivity rates at the PDV for those who acquired disease at least 14 days post dose 2 and were seronegative and PCR negative at baseline (primary endpoint Covid-19 cases): Full Analysis Set Population.

We assessed whether higher SARS-CoV-2 viral copies at the illness visit was associated with anti-N seropositivity at the PDV. The median viral copy number at the Day 1 illness visit was significantly higher in placebo recipients who were anti-N seropositive on the PDV (6.8 log_10_ copies/ml) than in placebo recipients who were anti-N seronegative on the PDV (2.2 log_10_ copies/ml) (p<.01). A similar result was seen in vaccine recipients, with analogous viral copy numbers of 6.1 log_10_ copies/ml and 2.4 log_10_ copies/ml, respectively (p<.01) (Figure 2A). Using logistic regression with terms for arm and viral copy number, an increase in illness visit viral copy number of 1 log_10_ nearly doubled the odds of anti-N seropositivity at the PDV (odds ratio 1.90 per 1-log_10_ increase; 95% confidence interval 1.59, 2.28). And yet, viral copies at the illness visit did not fully explain the large difference in PDV seropositivity between arms: for any given viral copy number, the odds of anti-N seropositivity were 13.67 times higher for the placebo arm than the vaccine arm (95% CI 5.17, 36.16). For example, a vaccine recipient with 2.0 log_10_ viral copies/ml on illness visit has an estimated probability of PDV anti-N seropositivity of 0.15, while for a placebo recipient with the same illness visit viral copy number, the estimated probability is 0.71 (Figure 2B). We also compared the overall prediction of PDV seropositivity using viral copies at the Day 1 illness visit versus the average over all illness visits; the latter measure is influenced by both magnitude and duration of viral replication. Viral copies at Day 1 illness performed better, with an area under the receiver operating characteristic curve of 0.84 versus 0.78 (p<0.05).

**Figure 2.**
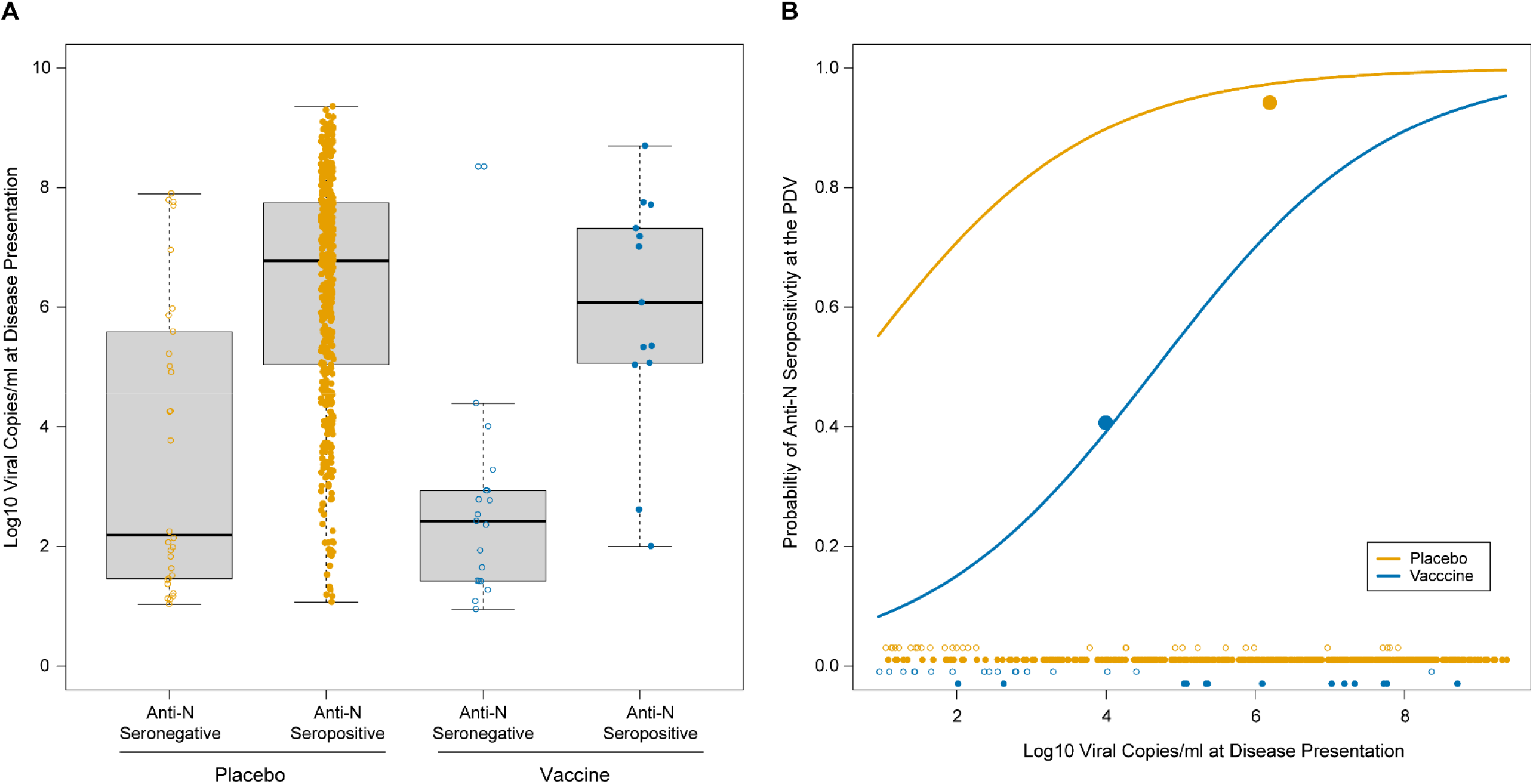
Relationship between viral copy number at disease presentation and anti-N serostatus at PDV by arm. A) Log10 viral copies/ml at disease presentation for placebo (orange circles) and vaccine (blue circles) arms by PDV anti-N serostatus. B) Predicted probability of PDV anti-N seropositivity by arm (orange, placebo; blue, vaccine). Closed and open circles denote anti-N seropositive and anti-N seronegative cases, respectively. The two large closed circles are plotted at the mean viral copy number and the mean PDV anti-N seropositivity rates by arm.

Table 2 presents the anti-N seropositivity rates at different study timepoints for infections detected at baseline, prior to receipt of study product. We found similar anti-N seropositivity rates between the two arms: anti-N seropositivity rates at Day 29 are 95.1% (270/284) and 96.1% (270/281) for the placebo and mRNA-1273 arms. These high rates are maintained through the PDV at a median of 149 days later). For baseline SARS-CoV-2 PCR positive, anti-N seronegative participants, the Day 29 anti-N seropositivity rates are 74.1% (20/27) and 73.3% (22/30) for the placebo and mRNA-1273 arms respectively, and are similar at the PDV with rates of 82.6% (19/23) and 71.0% (22/31), respectively. Within each arm, the Day 29 and PDV anti-N seropositivity rates are significantly lower for the baseline PCR positive participants compared to baseline anti-N seropositive participants (p<0.05 for each arm).

**Table 2.** Anti-N seropositivity rates at Day 29 and PDV for infections detected by serology or PCR at Day 1 (Baseline).

| Infection<br>Detection | Anti-N Serostatus at Day 29 |  |  |  | Anti-N serostatus at PDV |  |  |  | Median<br>Days<br>(IQR) |
| --- | --- | --- | --- | --- | --- | --- | --- | --- | --- |
|  | Placebo |  | mRNA |  | Placebo |  | mRNA |  |  |
|  | N | % Positive<br>(95% CI) | N | % Positive<br>(95% CI) | N | % Positive<br>(95% CI) | N | % Positive<br>(95% CI) |  |
| Baseline<br>Seropositive | 284 | 95.1%<br>(92.6%,<br>97.6%) | 281 | 96.1%<br>(93.8%,<br>98.4%) | 244 | 94.3%<br>(91.4%,<br>97.2%) | 260 | 93.8%<br>(90.9%,<br>96.7%) | 149.0<br>(76.0-236.0) |
| Baseline<br>PCR+* | 27 | 74.1%<br>(57.6%,<br>90.6%) | 30 | 73.3%<br>(57.5%,<br>89.1%) | 23 | 82.6%<br>(67.1%,<br>98.1%) | 31 | 71.0%<br>(55.0%,<br>87.0%) | 153.5<br>(86.0-<br>220.0) |

\*Also baseline seronegative.

Table 3 presents seropositivity rates for infections detected at Day 29 among baseline SARS-CoV-2 negative participants, which are post dose 1 and thus subject to partial vaccine effects. For anti-N seropositive participants on Day 29, the anti-N seropositivity rates at Day 57 are 86.9% (53/61) and 84.6% (33/39) for the placebo and mRNA-1273 arms. The rates are similar at the PDV, a median of 118 days later:84.3% (43/51) and 86.1% (31/36). These rates are not significantly different at Day 57 versus PDV or by arm (p>0.05 for all tests). For those who are newly RT-PCR positive and anti-N seronegative at Day 29, the analogous seropositive rates are 60.0% (18/30) and 38.5% (5/13) at Day 57 and 70.4% (19/27) and 50.0% (6/12) at the PDV. These rates are also not significantly different at Day 57 versus PDV or by arm (p>0.05). Consistent with the effects seen among baseline infections, the Day 57 and PDV anti-N seropositivity rates are significantly lower for Day 29 RT-PCR positive, anti-N seronegative participants compared to Day 29 anti-N seropositive participants (p<0.05).

**Table 3.**
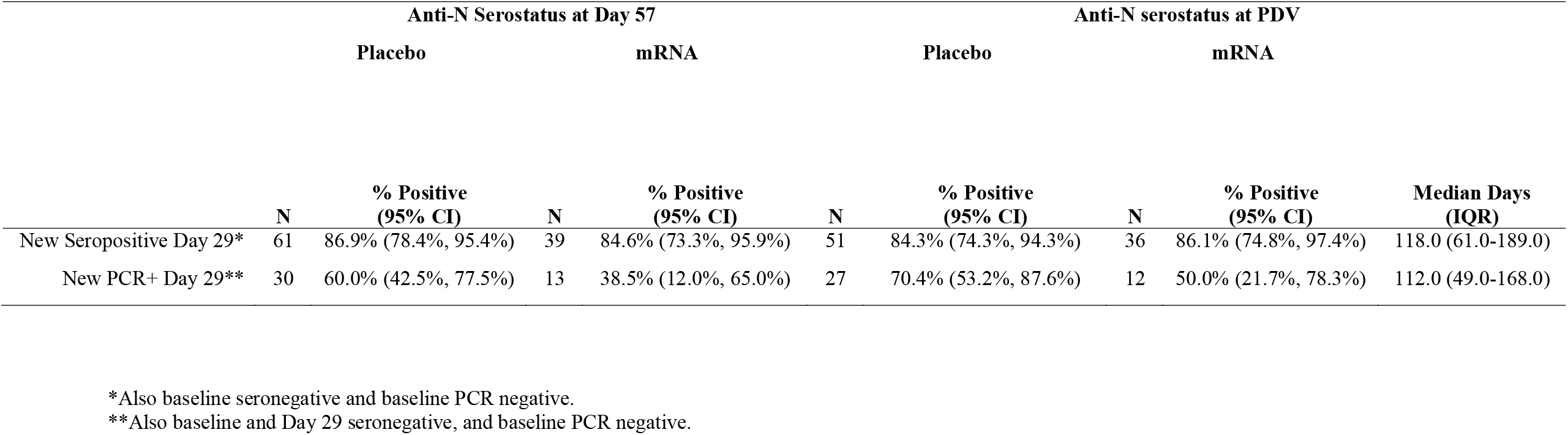
Anti-N seropositivity rates at Day 57 and PDV for infections detected by serology or PCR at Day 29 (dose 2).

## Discussion

These data show that, among the participants with PCR-confirmed Covid-19 disease, anti-N Ab seropositivity at a median of 53 days post diagnosis occurred in 40% of the mRNA-1273 vaccine recipients vs. 93% of the placebo recipients. While an increase in seroreversion cannot be ruled out, given the short time frame the more likely explanation is a vaccine-induced reduction in seroconversion. Anti-N seropositivity correlated with illness visit SARS-CoV-2 viral copy number, with each log increase in viral copy number nearly doubling the odds of anti-N seropositivity at the PDV. As the viral copy number on the day of the illness visit in mRNA-1273 vaccinated Covid-19 cases has been shown to be 100-fold lower than that in placebo recipient Covid-19 cases,^9^ the lower anti-N seropositivity in the mRNA-1273 recipients could be partly explained by their reduced exposure to N-antigen. However, strong vaccine effects remain; at 2.0 log_10_ copies/ml the predicted probability of seroconversion was 0.15 for vaccinated Covid-19-cases compared to 0.71 for placebo recipient Covid-19 cases. This may be due to a difference in the live virus replication between vaccine and placebo recipients, which cannot be differentiated by the RT-PCR test. Another potential explanation is that the vaccine has much larger effects on reducing replication outside the nose, as was shown in a study evaluating the mRNA-1273 vaccine against SARS-CoV-2 challenge in a non-human primate model.^11^ There may be other features of the initial course of infection that influence anti-N Ab seroconversion and that are affected by vaccination. Interestingly, the average viral copy number across post-Covid-19 illness visits was found to be a worse predictor of PDV seroconversion.

SARS-CoV-2 infection results in a wide spectrum of clinical outcomes from subclinical to fatal. Symptomatic Covid-19 of any severity and severe Covid-19 disease are of clear medical relevance. While subclinical SARS-CoV-2 infection is of less direct medical importance, it is well recognized that asymptomatic acquisition and carriage are important factors in household, institutional, and community wide transmission.^12-15^ Anti-N Ab seroconversion has been to date the major focus for defining subclinical and past Covid-19 detection, both on the individual and population levels.^16-18^

Our study has shown that anti-N seropositivity as evidence of previous SARS-CoV-2 infection is complex and may be subject to large vaccine effects. Multiple studies have reported that some fraction of PCR-confirmed SARS-CoV-2 infections are not accompanied by seroconversion; estimates range from 5% to 36%.^19-23^ While low rates of anti-N seroconversion in fully vaccinated (BNT162b2) hospital healthcare workers were observed in a large seroprevalence study in Ireland,^24^ our study provides the first evidence from a randomized, placebo-controlled trial with systematic surveillance for infection. This effect has consequences for interpretation of endpoints in vaccine trials, observational studies, serosurveys, and for monitoring and responding to the ongoing pandemic.

Even with frequent serosampling, serosurveys that rely on antibodies to the N protein may underestimate within-community transmission dynamics. A meta-analysis showed that of the studies that reported the assay used for diagnosing previous infection, 59% used N-targeting tests.^25^ In our study, the seroconversion rate was about 40% at a median of 53 days post-diagnosis for fully vaccinated individuals who developed Covid-19 disease. Following 1 dose, the seroconversion rates for those PCR positive at Day 29 were 38.5% at Day 57 and 50.0% at the PDV (median 112 days post-diagnosis); the seroconversion rates following 2 doses for those PCR positive could not be evaluated in our study but are likely lower. While anti-N Ab seroconversion is high for the unvaccinated, the proportion of people immunologically naïve to SARS-CoV-2 is diminishing as the global population acquires immunity through infection and/or vaccination.^26-28^ Hence, conclusions about the prevalence and incidence of SARS-CoV-2 infection based on serologic assays need to be weighed in the context of our results.

We found that participants with infection detected prior to vaccination, and those with infections diagnosed via serology prior to full vaccination, remained seropositive for the period of observation (to the PDV). Reductions in seroconversion rates were most evident in those who met primary endpoint Covid-19 case criteria, i.e. became ill <u>14 or more days</u> after full vaccination. Whether PCR positive, asymptomatic breakthrough infections seroconvert at reduced rates will require study in cohorts with systematic asymptomatic testing.

Covid-19 vaccine efficacy trials^5,29-31^ use anti-N Ab seroconversion as a secondary or exploratory endpoint to assess vaccine efficacy against asymptomatic SARS-CoV-2 infection, or as part of a secondary endpoint to assess vaccine efficacy against SARS-CoV-2 infection of any severity. As we found that infections in the placebo arm are about twice as likely to manifest through anti-N Ab seroconversion as those in the vaccine arm, our results suggest that caution is needed when interpreting vaccine estimates against such endpoints. One solution may be to define the endpoint of interest as an infection that is accompanied by seroconversion. This seroconversion endpoint would exclude ∼60% of infection cases and an unknown percentage of PCR positive cases in fully vaccinated individuals. The impact would likely be to reduce the overall sensitivity of the SARS-CoV-2 infection case definition. It is also unknown whether non-seroconverting infections post vaccination are less likely to transmit to others. An additional approach that may reflect transmission potential would be to contrast the mean viral loads over all individuals at a single point in time (or over the course of the acute illness), a metric known as vaccine efficacy on prevalent viral load.^32,33^ While the lack of anti-N seroconversion may indicate that the breakthrough infection was immunologically ‘silent’, other immunological markers may be useful to determine these infections. These include SARS-CoV-2-specific cellular responses against the N and M proteins.^34^ These tests are at present expensive and often require special sampling and processing techniques that make it difficult to implement them for large scale seroprevalence studies, but would be important to explore in a research setting.

This analysis has a few limitations. First, our participants received exclusively mRNA-1273 vaccine. It is unknown if vaccination with BNT162b2 or Ad26.COV2.S interferes with anti-N seropositivity post infection to the same magnitude as mRNA-1273. In a study of 31 individuals, the majority of whom received BNT162b2, and then became infected with SARS-CoV-2, anti-N Ab were detected in 68% at 5 weeks post infection using ELISA.^35^ Second, we only used the Elecsys assay, which has some of the highest sensitivity and specificity data, and it is unknown if other immunoassays will have different sensitivity at detecting recent or past infections in vaccinated persons. Third, our sample size is small with 52 fully-vaccinated persons with breakthrough Covid-19 assessed for seroconversion a median of 53 days post-diagnosis; thus transient anti-N seropositivity events could go undetected. However, the close follow up and uniformity in obtaining the swabs and serum samples increase confidence in and interpretability of our findings. Fourth, SARS-CoV-2 infections post vaccination that are caused by the delta and omicron variants have been associated with higher viral copy number upon diagnosis and more rapid clearance.^36,37^ Whether breakthrough infections due to these variants is associated with different rates of seroconversion cannot be ascertained using these data. Finally, this analysis was not addressing a pre-specified objective of the COVE study. While the statistical analysis plan was pre-specified, the COVE study was not designed to assess the sensitivity/specificity of the Elecsys assay in vaccinees versus placebo recipients.

In summary, these data support that there are potential assay limitations in detecting anti-N Abs in recently mRNA-1273-vaccinated individuals. Determining SARS-CoV-2 infections at a population level via serosurveillance in the era of Covid-19 vaccination coverage requires further research on detection of recent or remote SARS-CoV-2 infection in vaccinated individuals, and vaccination status should be taken into account when interpreting seroprevalence and seropositivity data.

## Data Availability

As the trial is ongoing, access to participant-level data and supporting clinical documents with qualified external researchers may be available upon request and is subject to review once the trial is complete.

## Ethics

The mRNA-1273-P301 study is being conducted in accordance with the International Council for Harmonisation of Technical Requirements for Pharmaceuticals for Human Use, Good Clinical Practice guidelines, and applicable government regulations. The Central Institutional Review Board approved the mRNA-1273-P301 protocol and the consent forms. All participants provided written informed consent before enrollment. Central IRB services for the mRNA-1273-P301 study were provided by Advarra, Inc., 6100 Merriweather Dr., Suite 600, Columbia, MD 21044.

## Acknowledgments

We thank the volunteers who participated in the COVE trial. We also thank Lindsay Carpp for assistance with scientific writing and technical editing.

---

**eFigure 1.**
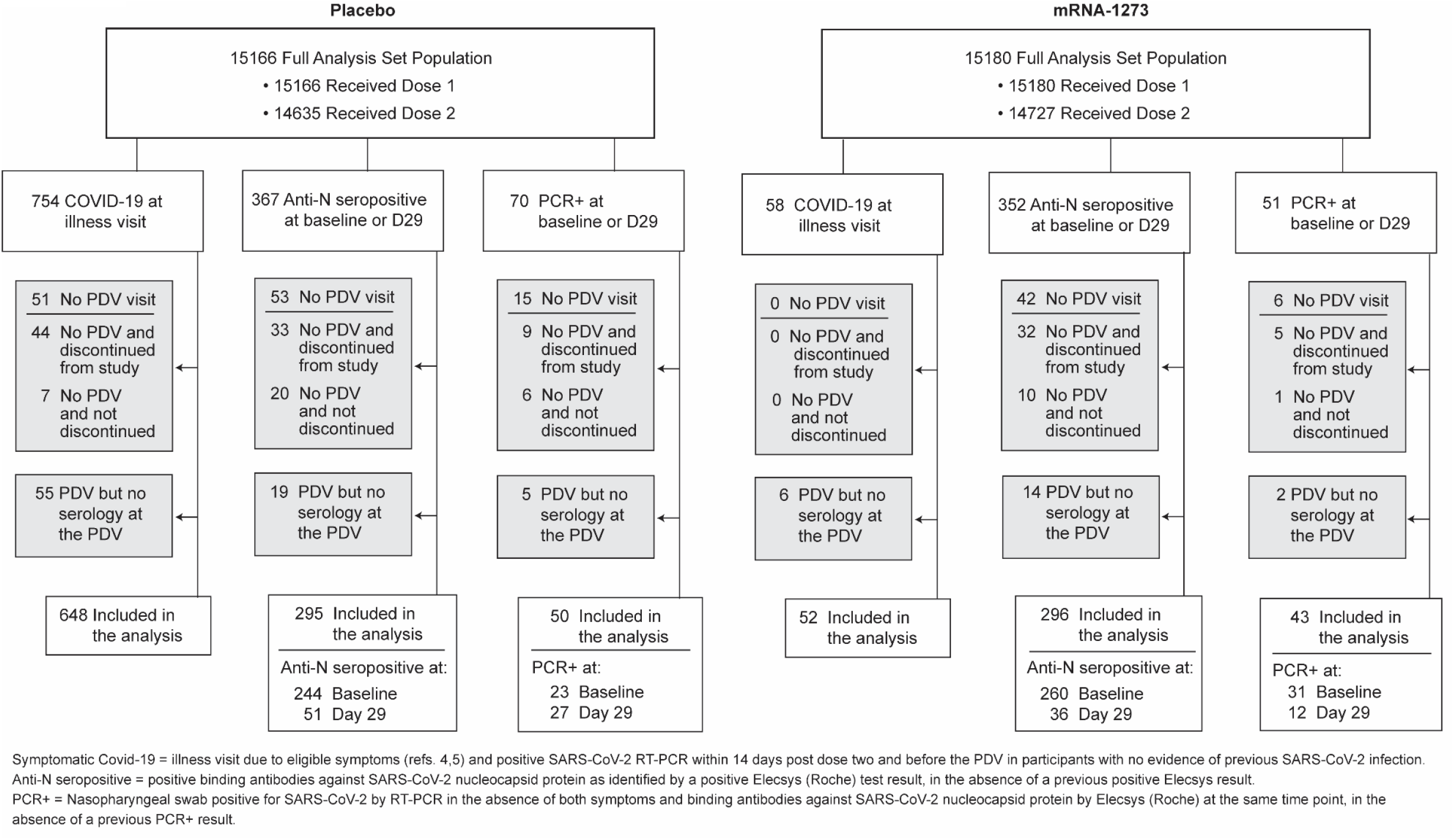
Flow of participants with detected SARS-CoV-2 infection from enrollment through to inclusion in the analysis. *Six participants with major protocol deviations and two participants who were randomized twice were excluded from the full analysis set. D29 and D57 denote the Day 29 and the Day 57 study visits, respectively. PDV, participant decision visit. Data cutoff date: March 26, 2021.

**eTable 1.** Participant baseline characteristics overall and by arm in the population in whom post-baseline infections were first detected at an illness visit.

|  | Full Analysis<br>Set Population<br>Total<br>N=30346 | Post-Baseline Infections First Detected<br>at an Illness Visit |  |
| --- | --- | --- | --- |
|  |  | Placebo<br>N=754 | mRNA-1273<br>N=58 |
| **Sex - no. of participants (%)** |  |  |  |
| Female | 14372 (47.4) | 372 (49.3) | 27 (46.6) |
| Male | 15974 (52.6) | 382 (50.7) | 31 (53.4) |
| **Age in yrs – Mean (Range)** | 51.4 (18.0-95.0) | 47.8 (18.0-87.0) | 48.5 (24.0-74.0) |
| **Hispanic or Latino ethnicity- no. of participants (%)** |  |  |  |
| Hispanic or Latino | 6230 (20.5) | 179 (23.7) | 10 (17.2) |
| Not Hispanic or Latino | 23838 (78.6) | 571 (75.7) | 48 (82.8) |
| Not reported and unknown | 278 (0.9) | 4 (0.5) | 0 (0.0) |
| **Race or ethnic group - no. of participants (%)** |  |  |  |
| American Indian or Alaska Native | 234 (0.8) | 5 (0.7) | 0 (0.0) |
| Asian | 1395 (4.6) | 29 (3.8) | 1 (1.7) |
| Black or African American | 3098 (10.2) | 41 (5.4) | 4 (6.9) |
| Multiple | 638 (2.1) | 9 (1.2) | 1 (1.7) |
| Native Hawaiian or Other Pacific | 68 (0.2) | 0 (0.0) | 0 (0.0) |
| Not reported and unknown | 288 (0.9) | 11 (1.5) | 0 (0.0) |
| Other | 593 (2.0) | 20 (2.7) | 1 (1.7) |
| White | 24032 (79.2) | 639 (84.7) | 51 (87.9) |
| **Age category and risk for severe Covid-19 - no. of participants (%)** |  |  |  |
| >=18 and <65 yr, not at Risk | 17770 (58.6) | 489 (64.9) | 35 (60.3) |
| >=18 and <65 yr, at Risk | 5065 (16.7) | 165 (21.9) | 14 (24.1) |
| >=65 Years | 7511 (24.8) | 100 (13.3) | 9 (15.5) |
| **Risk factor for severe Covid-19- no. of participants (%)** |  |  |  |
| Chronic Lung Disease | 1461 (4.8) | 32 (4.2) | 4 (6.9) |
| Significant Cardiac Disease | 1504 (5.0) | 30 (4.0) | 4 (6.9) |
| Severe Obesity | 2128 (7.0) | 77 (10.2) | 8 (13.8) |
| Diabetes | 2917 (9.6) | 72 (9.5) | 3 (5.2) |
| Liver Disease | 200 (0.7) | 5 (0.7) | 1 (1.7) |
| Human Immunodeficiency Virus Infection | 185 (0.6) | 4 (0.5) | 0 (0.0) |

**eTable 2.** Participant baseline characteristics overall and by arm in the population in whom infection was first detected at baseline by anti-N seropositivity or PCR positivity.

|  | Full Analysis Set<br>Population<br>Total<br>N=30346 | Infections Detected at Baseline By: |  |  |  |
| --- | --- | --- | --- | --- | --- |
|  |  | Anti-N Seropositivity |  | PCR+ |  |
|  |  | Placebo<br>N=303 | mRNA-1273<br>N=309 | Placebo<br>N=34 | mRNA-1273<br>N=38 |
| **Sex - no. of participants (%)** |  |  |  |  |  |
| Female | 14372 (47.4) | 142 (46.9) | 135 (43.7) | 14 (41.2) | 17 (44.7) |
| Male | 15974 (52.6) | 161 (53.1) | 174 (56.3) | 20 (58.8) | 21 (55.3) |
| **Age in yrs – Mean (Range)** | 51.4 (18.0-95.0) | 45.0 (18.0 - 83.0) | 44.3 (18.0 - 4.0) | 45.7 (19.0-74.0) | 42.4 (20.0-84.0) |
| **Hispanic or Latino ethnicity- no. of participants (%)** |  |  |  |  |  |
| Hispanic or Latino | 6230 (20.5) | 140 (46.2) | 139 (45.0) | 11 (32.4) | 7 (18.4) |
| Not Hispanic or Latino | 23838 (78.6) | 161 (53.1) | 168 (54.4) | 23 (67.6) | 31 (81.6) |
| Not reported and unknown | 278 (0.9) | 2 (0.7) | 2 (0.6) | 0 (0.0) | 0 (0.0) |
| **Race or ethnic group - no. of participants (%)** |  |  |  |  |  |
| American Indian or Alaska Native | 234 (0.8) | 2 (0.7) | 0 (0.0) | 0 (0.0) | 0 (0.0) |
| Asian | 1395 (4.6) | 4 (1.3) | 10 (3.2) | 1 (2.9) | 0 (0.0) |
| Black or African American | 3098 (10.2) | 88 (29.0) | 91 (29.4) | 2 (5.9) | 8 (21.1) |
| Multiple | 638 (2.1) | 2 (0.7) | 5 (1.6) | 0 (0.0) | 0 (0.0) |
| Native Hawaiian or Other Pacific | 68 (0.2) | 0 (0.0) | 0 (0.0) | 1 (2.9) | 0 (0.0) |
| Not reported and unknown | 288 (0.9) | 4 (1.3) | 5 (1.6) | 0 (0.0) | 0 (0.0) |
| Other | 593 (2.0) | 7 (2.3) | 9 (2.9) | 1 (2.9) | 0 (0.0) |
| White | 24032 (79.2) | 196 (64.7) | 189 (61.2) | 29 (85.3) | 30 (78.9) |
| **Age category and risk for severe Covid-19 - no. of participants (%)** |  |  |  |  |  |
| >=18 and <65 yr, not at Risk | 17770 (58.6) | 213 (70.3) | 233 (75.4) | 23 (67.6) | 30 (78.9) |
| >=18 and <65 yr, at Risk | 5065 (16.7) | 61 (20.1) | 43 (13.9) | 6 (17.6) | 5 (13.2) |
| >=65 Years | 7511 (24.8) | 29 (9.6) | 33 (10.7) | 5 (14.7) | 3 (7.9) |
| **Risk factor for severe Covid-19- no. of participants (%)** |  |  |  |  |  |
| Chronic Lung Disease | 1461 (4.8) | 11 (3.6) | 6 (1.9) | 1 (2.9) | 2 (5.3) |
| Significant Cardiac Disease | 1504 (5.0) | 10 (3.3) | 10 (3.2) | 2 (5.9) | 1 (2.6) |
| Severe Obesity | 2128 (7.0) | 22 (7.3) | 24 (7.8) | 3 (8.8) | 3 (7.9) |
| Diabetes | 2917 (9.6) | 23 (7.6) | 20 (6.5) | 1 (2.9) | 4 (10.5) |
| Liver Disease | 200 (0.7) | 1 (0.3) | 1 (0.3) | 0 (0.0) | 0 (0.0) |
| Human Immunodeficiency Virus Infection | 185 (0.6) | 7 (2.3) | 4 (1.3) | 0 (0.0) | 0 (0.0) |

**eTable 3.** Participant baseline characteristics overall and by arm in the population in whom post-baseline infections were first detected by anti-N seropositivity or by PCR.

|  | Full Analysis<br>Set Population<br>Total<br>N=30346 | Post-Baseline Infections First Detected By: |  |  |  |
| --- | --- | --- | --- | --- | --- |
|  |  | Anti-N Seropositivity (Day 29)<br>Placebo<br>N=64 | mRNA-1273<br>N=43 | PCR+ (Day 29)<br>Placebo<br>N=36 | mRNA-1273<br>N=13 |
| **Sex - no. of participants (%)** |  |  |  |  |  |
| Female | 14372 (47.4) | 33 (51.6) | 14 (32.6) | 15 (41.7) | 7 (53.8) |
| Male | 15974 (52.6) | 31 (48.4) | 29 (67.4) | 21 (58.3) | 6 (46.2) |
| **Age in yrs – Mean (Range)** | 51.4 (18.0-95.0) | 48.0 (19.0-76.0) | 41.9 (20.0-70.0) | 46.3 (18.0-77.0) | 50.6 (23.0-69.0) |
| **Hispanic or Latino ethnicity-**<br>**no. of participants (%)** |  |  |  |  |  |
| Hispanic or Latino | 6230 (20.5) | 19 (29.7) | 8 (18.6) | 11 (30.6) | 2 (15.4) |
| Not Hispanic or Latino | 23838 (78.6) | 44 (68.8) | 35 (81.4) | 25 (69.4) | 11 (84.6) |
| Not reported and unknown | 278 (0.9) | 1 (1.6) | 0 (0.0) | 0 (0.0) | 0 (0.0) |
| **Race or ethnic group - no. of**<br>**participants (%)** |  |  |  |  |  |
| American Indian or Alaska Native | 234 (0.8) | 0 (0.0) | 0 (0.0) | 1 (2.8) | 0 (0.0) |
| Asian | 1395 (4.6) | 2 (3.1) | 0 (0.0) | 1 (2.8) | 0 (0.0) |
| Black or African American | 3098 (10.2) | 9 (14.1) | 13 (30.2) | 5 (13.9) | 2 (15.4) |
| Multiple | 638 (2.1) | 0 (0.0) | 2 (4.7) | 2 (5.6) | 0 (0.0) |
| Native Hawaiian or Other Pacific | 68 (0.2) | 0 (0.0) | 0 (0.0) | 0 (0.0) | 0 (0.0) |
| Not reported and unknown | 288 (0.9) | 2 (3.1) | 0 (0.0) | 0 (0.0) | 0 (0.0) |
| Other | 593 (2.0) | 2 (3.1) | 1 (2.3) | 0 (0.0) | 1 (7.7) |
| White | 24032 (79.2) | 49 (76.6) | 27 (62.8) | 27 (75.0) | 10 (76.9) |
| **Age category and risk for**<br>**severe Covid-19 - no. of**<br>**participants (%)** |  |  |  |  |  |
| >=18 and <65 yr, not at Risk | 17770 (58.6) | 41 (64.1) | 30 (69.8) | 23 (63.9) | 7 (53.8) |
| >=18 and <65 yr, at Risk | 5065 (16.7) | 11 (17.2) | 9 (20.9) | 6 (16.7) | 3 (23.1) |
| >=65 Years | 7511 (24.8) | 12 (18.8) | 4 (9.3) | 7 (19.4) | 3 (23.1) |
| **Risk factor for severe Covid-**<br>**19- no. of participants (%)** |  |  |  |  |  |
| Chronic Lung Disease | 1461 (4.8) | 2 (3.1) | 1 (2.3) | 0 (0.0) | 1 (7.7) |
| Significant Cardiac Disease | 1504 (5.0) | 3 (4.7) | 1 (2.3) | 0 (0.0) | 1 (7.7) |
| Severe Obesity | 2128 (7.0) | 3 (4.7) | 6 (14.0) | 3 (8.3) | 3 (23.1) |
| Diabetes | 2917 (9.6) | 5 (7.8) | 2 (4.7) | 5 (13.9) | 1 (7.7) |
| Liver Disease | 200 (0.7) | 0 (0.0) | 0 (0.0) | 0 (0.0) | 0 (0.0) |
| Human Immunodeficiency Virus<br>Infection | 185 (0.6) | 0 (0.0) | 0 (0.0) | 0 (0.0) | 0 (0.0) |

